# A Bayesian analysis of the total number of cases of the COVID 19 when only a few data is available. A case study in the state of Goias, Brazil

**DOI:** 10.1101/2020.04.19.20071852

**Authors:** Renato Rodrigues Silva, Wisley Donizetti Velasco, Wanderson da Silva Marques, Carlos Augusto Gonçalves Tibiriçá

## Abstract

The outbreak of COVID 19 has been provoking several problems to the health system around the world. One of the concerning is the crash of the health system due to the increasing demand suddenly. To avoid it, knowing the total number and daily new cases is crucial. In this study, we fitted curves growth models using a Bayesian approach. We extracted information obtained from some countries to build the prior distribution of the model. The total number of cases of the COVID 19 in the state of Goias was analyzed. Results from analysis indicated that the date of the outbreak peak is between 51 and 68 days after the beginning. Moreover, the total number of cases is around 3180 cases. The analysis did not take into consideration possibles changes in government control measures. We hope this study can provide some valuable information to public health management.

## 1 Introduction

The outbreak of COVID 19 has been provoking several problems to the health system around the world. The World Health Organization classified the COVID 19 outbreak as pandemic on March 11, 2020 ([19]; [20]). Brazil confirmed the first case of COVID 19 on 26, February 2020 [1].

Since then, the outbreak has gotten attention from researchers, media, authorities, and society in general ([14];[19]).

During an infectious disease outbreak, one of the concerning is the crash of the health system due to the increasing demand suddenly. In this context, knowing the total number and daily new cases is crucial.

In the literature, there are many statistical and mathematical methods to solve this problem. Compartment models ([17]; [22]), partially observed Markov process [9], temporal network analysis [13], growth curves ([16]; [24]; [25]) and others.

In this study, we fitted curves growth models to the dataset. Predict the values of a time series, assuming a Gompertz or logistic trend, is not a new idea [23]. [2] fitted the Gompertz curve to predict the total number of cases of the COVID 19 in Iran.

Here we used Bayesian inference. The main advantage of this approach is to include other sources of the information besides the dataset itself. For this particular case, we extracted information obtained from some countries to build the prior distribution of the model. We analyzed the data from the state of Goias, Brazil, as an example.

## 2 Material and Methods

The data of the countries analyzed in this study can be found at [26] and the total number and daily new cases of the COVID 19 in the state of Goias are available at [8].

The main objective of the study is to make predictions of the total number of cases of COVID 19 in the state of Goias, Brazil. For this purpose, we fit a Gompertz and Logistic’s non-linear regression [25] to the data using the Bayesian inference approach [11]. However, we are at the beginning of the outbreak and therefore have a few data available. To figure out this problem, we included prior information from results obtained of the fitting frequentist non-linear to data of other countries.

Initially, we fitted a Gompertz and Logistic non-linear regression model to data of total number of cases of the COVID 19 for some countries, such as China, Germany, Italy, Philippines, South Korea, Spain, Taiwan, Thailand and the United States of America. We have chosen these countries because all of them are in the advanced stage of the COVID 19 outbreak, and there is significant variability among them.

The Gompertz non linear regression model can be expressed as

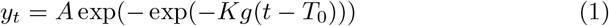

where *y*_*t*_ is the total number of cases of COVID 19 observed at *t* − *th* time, *A* is the upper asymptote that represents the total case at the end of the outbreak, *K*_*g*_ is the growth-rate coefficient and *T*_0_ represent time at inflection, or the time when will occur the maximum daily new cases.

The Logistic non linear regression model can be described mathematically by

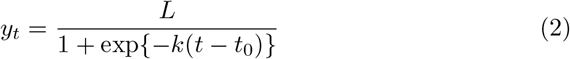

where *y*_*t*_ is the total cases of COVID 19 observed at *t* − *th* time, *L* is the upper asymptote, *k* is the growth-rate coefficient and *t*_0_ represent time at inflection.

For the data set of these countries, we use the maximum likelihood estimator because we have a quantity enough of data. Non-linear regression was fitted using nlsLM function of R package minpack.lm [10] of R statistical software [21]. Model selection between Gompertz and Logistic was done via Akaike Information Criterion [3].

From the results of the non-linear models fitted by maximum likelihood, we build the prior distribution of the parameters of the Bayesian non-linear models used to estimate the total number of cases of Goias state.

For each country, we have a correspondent model and their maximum likelihood estimates for the parameters of Gompertz’s model (*A, K*_*g*_, *T*_0_), and of the logistic model(*L, k, t*_0_), respectively.

The idea is to use each maximum likelihood estimate as a hypothesis and use them as to the prior distribution. In this step, we are inferring the resemblance between the Goias state curve and curve of other countries.

For parameters *A* and *L*, we have to take into considerations that each country has a different population and different than the Goiás state. Hence, we estimate the prevalence of the disease for an i-th country from expression *A*_*i*_ divided by *N*_*i*_, where *N*_*i*_ is the population of that country. After that, we multiplied the prevalence by the population of Goiás state. This number is equivalent to the total number of cases that would occur in Goiás, whether the hypothesis is true.

For example, let’s suppose we have 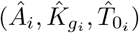 to be a Gompertz’s model maximum likelihood estimates for i-th country.

In this case, the prior can be expressed as

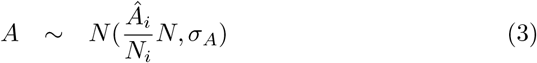

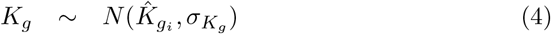

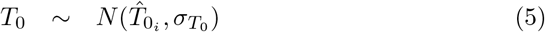

where 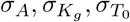 are standard deviation of prior distribution. We used the standard error of maximum likelihood estimates as standard deviation of prior distribution.

Parameters inference was done via Gibbs Sampling algorithm [11] We used burn in equal to 5000 and number of iterations equal to 10000.

We did the sensibility analysis of prior distribution and comparison between models the using Watanabe – Akaike information criterion (WAIC) [4]. All Bayesian computations were done using the bmrs R package ([5]; [6]).

After sensibility analysis, we set an additional model building a prior distribution based on subjective evaluation of values of all maximum likelihood estimates obtained for each country.

For the variable total number of cases of COVID 19, we chose the model with the smallest WAIC, and then, we replaced the standard deviation of upper asymptote and time inflection by a value much higher with the objective to give freedom to parameters learn with data. We proceed that way because there is much uncertainty about the total number of cases of COVID 19. In this stage of the outbreak, we do not have any specific value of reference for upper asymptote and time inflection.

## 3 Results

Figure 1 shows bar plots for the daily number of new cases and scatter plots for the total number of cases of COVID 19 for some countries around the world.

**Figure 1:**
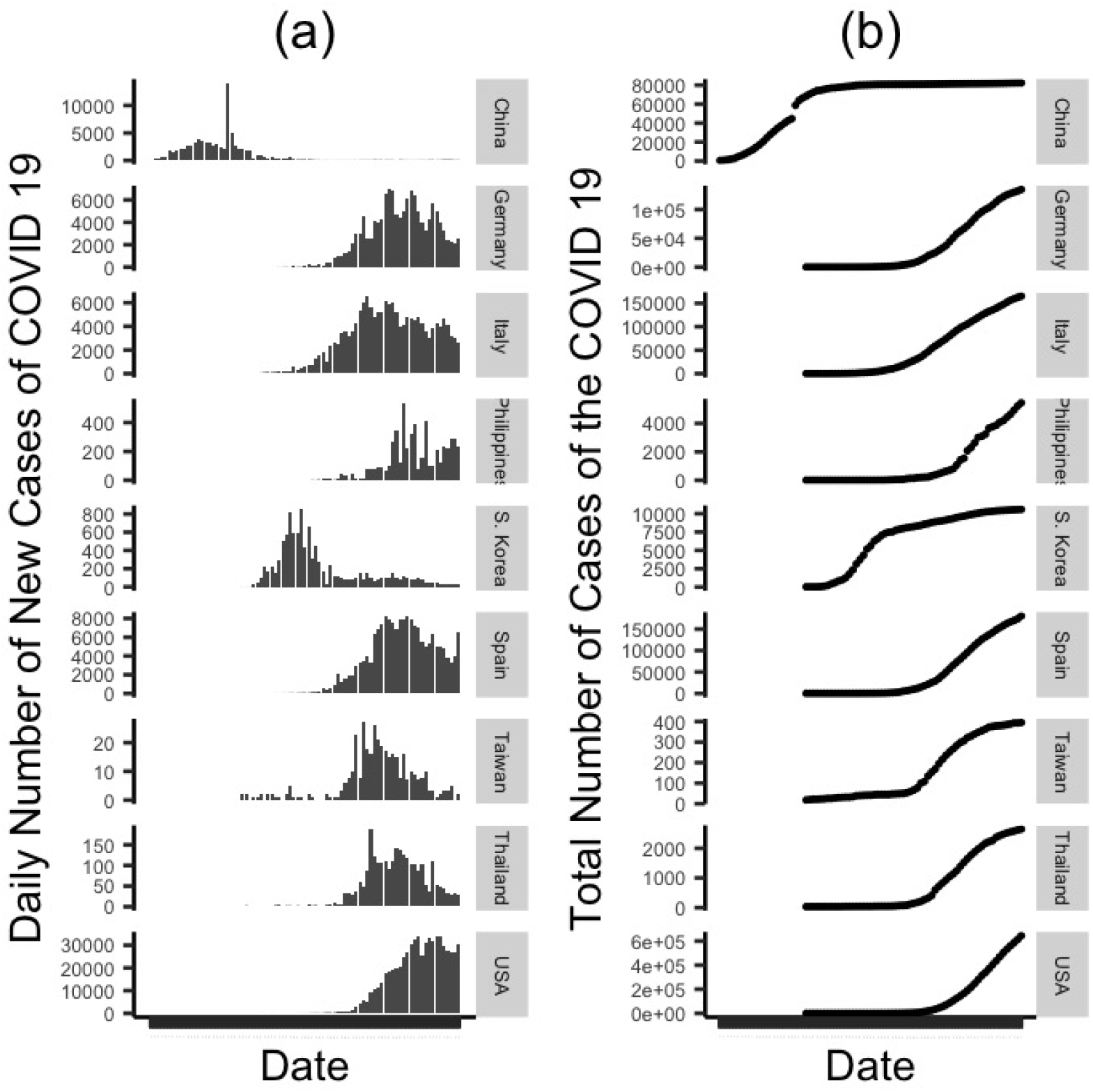
Bar plots for daily number of new cases(a) and scatter plots for total number of cases of the COVID 19 for each country analyzed (b)

Except for the United States of America, all countries analyzed possibly have already crossed the inflection point of the curve. China is the most advanced country in the pandemic. South Korea’s curve is singular. After the curve has been reaching out of the plateau, the total number of cases of COVID 19 still grows, indicating a possible “second wave” of the pandemic.

Table 1 and 2 shows the maximum likelihood estimates for the parameters of Gompertz and logistic model, respectively.

**Table 1:**
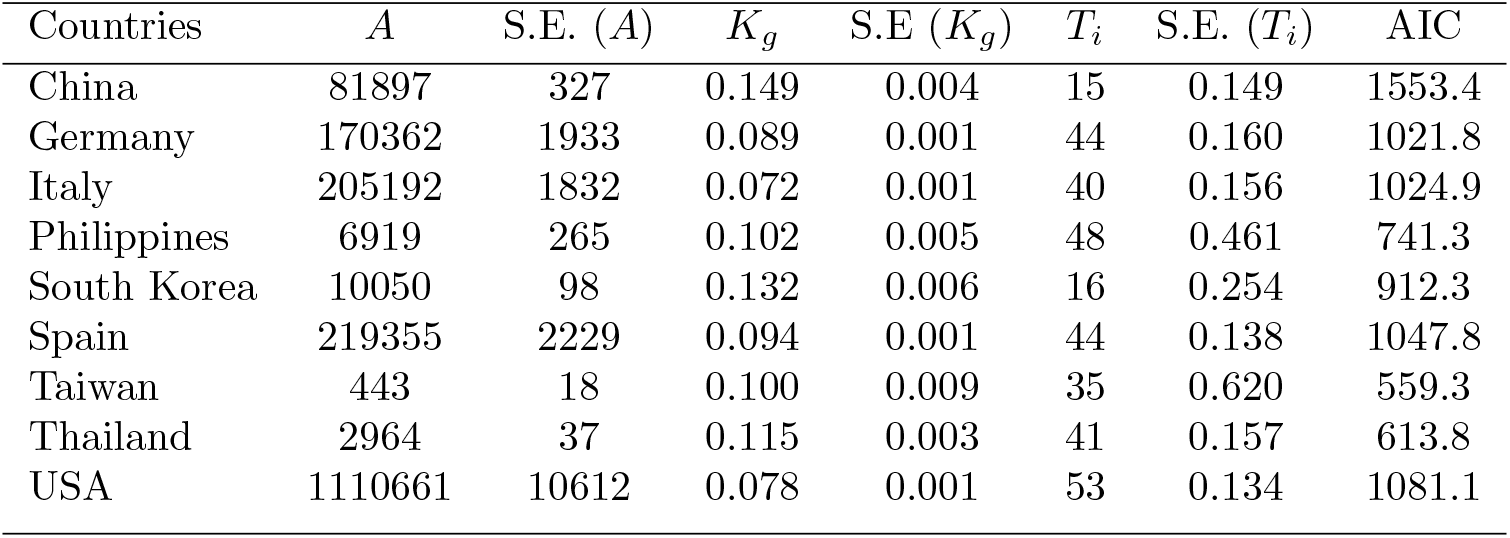
Maximum likelihood estimates and their respective standard errors of the parameters of the Gompertz model fitted to the total number of cases. The parameters are upper asymptote (*A*), growth rate (*K*_*g*_) and (*T*_*i*_) time for inflection

**Table 2:** Maximum likelihood estimates and their respective standard errors of the parameters of the logistic model fitted to the total number of cases. The parameters are upper asymptote (*L*), growth rate (*k*) and (*t*_0_) time for inflection

| Countries | $L$ | S.E. ($L$) | $k$ | S.E ($k$) | $t_0$ | S.E. ($t_0$) | AIC |
| --- | --- | --- | --- | --- | --- | --- | --- |
| China | 81260 | 213 | 0.222 | 0.004 | 18 | 0.103 | 1491.240 |
| Germany | 141176 | 1166 | 0.178 | 0.003 | 45 | 0.140 | 1064.7 |
| Italy | 169567 | 2025 | 0.143 | 0.003 | 42 | 0.250 | 1141.7 |
| Philippines | 5541 | 112 | 0.208 | 0.007 | 49 | 0.284 | 750.589 |
| South Korea | 9777 | 113 | 0.207 | 0.013 | 18 | 0.353 | 950.8 |
| Spain | 183703 | 1531 | 0.184 | 0.003 | 45 | 0.139 | 1103.2 |
| Taiwan | 413 | 8 | 0.159 | 0.009 | 38 | 0.468 | 526.5 |
| Thailand | 2675 | 21 | 0.202 | 0.004 | 43 | 0.139 | 621.5 |
| USA | 742189 | 10111 | 0.189 | 0.003 | 51 | 0.185 | 1234.5 |

Gompertz’s model provided higher values of upper asymptote and their respective standard errors than the logistic model. On the other hand, the estimates of the time inflection are pretty much the same for both models.

We can explain the difference between estimates of the upper asymptote, analyzing the shape of each curve and the dataset. Firstly, the abscissa of the point of the inflection of the logistic model corresponds to the half between two asymptotes, i. e. 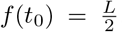. On the other hand, the abscissa of the point of the inflection of the Gompertz’s model is slightly smaller, 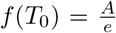 (references). Now, let’s suppose we have the same point of inflection for both curve, i.e, *f* (*t*_0_) = *f* (*T*_0_) = *f*. Thus, we have *A* = *ef* and *L* = 2*f*, which implies *A > L*. Furthermore, the COVID19 outbreaks is not in the plateau for most of the countries. Therefore, there is much uncertainty about the upper asymptotes of the curve, which explains the high values of standard errors.

China and South Korea reached out to the plateau faster than remainings countries. Taiwan has the least total number of cases followed by Thailand. We know that the outbreak is just the beginning. However, the COVID 19 outbreak control in Taiwan can be considered as a success case. Possible explanations for that can be effective containment and or extensive COVID-19 screening, effective patient triage, and contact tracing ([19]; [12]).

In contrast, the United States of America, Spain, Italy, and Germany are the top four countries in the total number of cases of COVID 19. We have to analyze the results from the United States of America carefully, especially the estimates of upper asymptotes. Once that the standard errors are quite high. However, we can state that likely the USA will have the largest total number of cases of COVID 19. Additionally, we have to be caution to compare Germany, Italy, and Spain. Because our model did not take consideration the sub notification, so the effects such as robust quarantine, early and complete identification and investigation of all contacts, social isolation, good habits about hygiene etiquette are confounded by sub notification and delay or mistakes in the recorded of the cases [27].

Results from AIC values (tables 1 and 2) revealed that the logistic model was the best only to data of China and Taiwan. [15] also analyzed data from China using Gompertz and Logistic Model. They also concluded that Logistic was the best model We already expected that Gompertz would be the best model for the majority of the models because the logistic curve has radial symmetry concerning the point of the inflection, which is rare for the spreading of diseases [25].

Figure 2 displays total and new cases of COVID 19 with their respective curves fitted. The lack of randomness of the total cases allows the fitting of the model with a sum of residuals quite low. We obtained the curve of the new cases computing the difference between the total cases observed at time t and the previous one.

**Figure 2:**
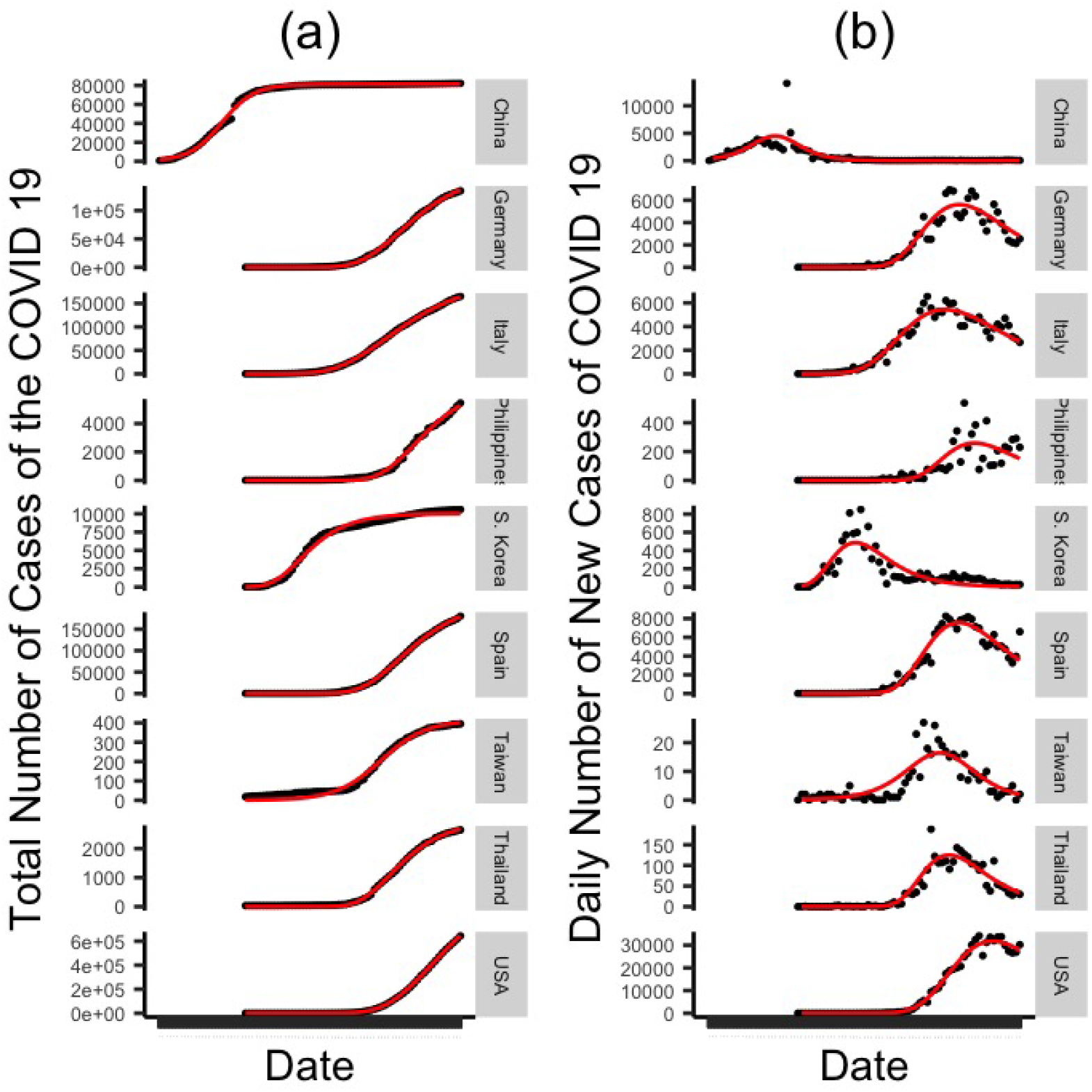
Total number of cases of COVID 19 with their respective curve fitted by non-linear regression (a) and Daily number of new cases of COVID 19 and their fitted curve (b) for each country analyzed

We do not have any intention to do a complete global analysis of COVID 19 outbreaks. Definitively, it is not the aim of our study. However, we aim understand what is happening in countries analyzed to extract priors information for our Bayesian models.

The interpretation of the prior distribution as follows. For instance, the use of MLE from China or the South Korea data set means to state that Goias states have a fast growth of outbreak and low total cases at the end. Philippines’ prior distribution means slow growth and moderate total cases. And for the United States of America, slow growth and high total cases. The maximum likelihood estimates for upper asymptote were not used directly. As described in the method section, an equivalent total number was computed first. This step of statistical analysis try to answer the following question: Which country the Goias COVID 19 outbreak curve resembles?

Table 3 shows the Watanabe Akaike information criterion computed from the fitting of Bayesian Gompertz and logistic to data of the total number of cases COVID 19 in the State of Goias, Brazil. Priors distributions based on maximum likelihood estimates from data of other countries

**Table 3:** Watanabe Akaike information criterion computed for Bayesian models for the total number of cases COVID 19 in the State of Goias using different priors distributions based on maximum likelihood estimated from data of other countries.

| Prior | WAIC (Gompertz) | WAIC (logistic) |
| --- | --- | --- |
| MLE China | 422.9 | 416.8 |
| MLE Germany | 461.4 | 480.6 |
| MLE Italy | 598.8 | 596.2 |
| MLE Philippines | 425.2 | 423.4 |
| MLE South Korea | 535.4 | 536.9 |
| MLE Spain | 525.8 | 548.7 |
| MLE Taiwan | 418.2 | 417.3 |
| MLE Thailand | 422.4 | 425.9 |
| MLE USA | 394.7 | 355.5 |

According to the WAIC criterion, the Goias COVID 19 outbreak curve is closer to the USA curve. It does not mean that the Goias state will have the total number of cases of COVID 19 approximately equal to the USA. Or even the same prevalence rate. It means there is a resemblance between two curves. Furthermore, the US dataset maximum likelihood estimates used as prior provides the predicted values more accurate among all prior distributions used so far.

The resemblances between the two curves allow us to make some comparisons. As much the USA as the Goias state did not do massive COVID 19 screening in their respective populations. There is no contact tracing of the infected in both locations. It was not done any prevention on airports, bus stations, and roads at the beginning of the outbreak.

Due to the high degree of uncertainty about upper asymptote, we decide to carry out an extra model with flat priors from the USA data set maximum likelihood estimates. Table 4 shows the results of the Bayesian inference for Gompertz with priors defined by *A ∼ N* (23000, 500000), *K*_*g*_ *∼ N* (0.08, 0.01), and *T*_0_ *∼ N* (53, 5); and logistic models with *L ∼ N* (15756, 500000), *k ∼ N* (0.189, 0.003), and *t*_0_ *∼* (51, 7).

**Table 4:**
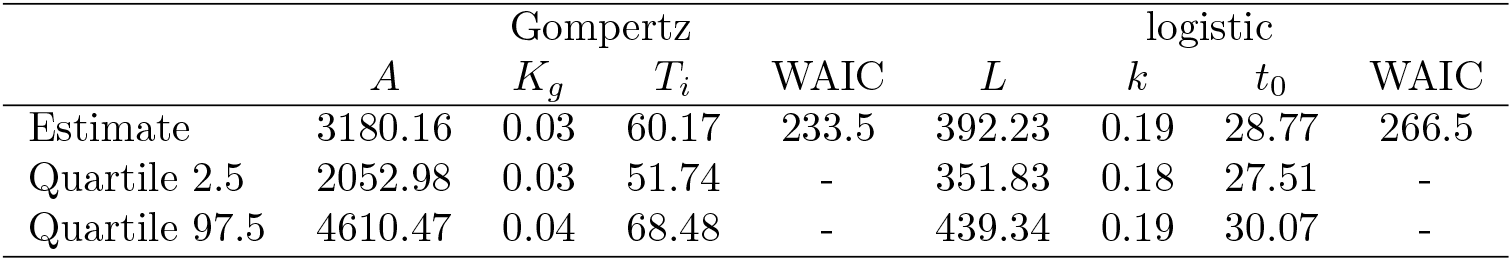
Results of the Bayesian inference for Gompertz and logistic model fitted to Goias’s data, with flat prior for upper asymptotes

The Gompertz model was the best using WAIC. The estimate of the data of the outbreak peak is 60 days after the beginning. The 95 % credibility interval indicates that the outbreak data can be ranges between 51 and 68 days after the outbreak initial. The estimate of the plateau is around 3180 cases, which corresponds to a prevalence rate of 4.53 per 10000 inhabitants. Figures 4 and 5 show the forecast for the total number and new daily cases.

**Figure 3:**
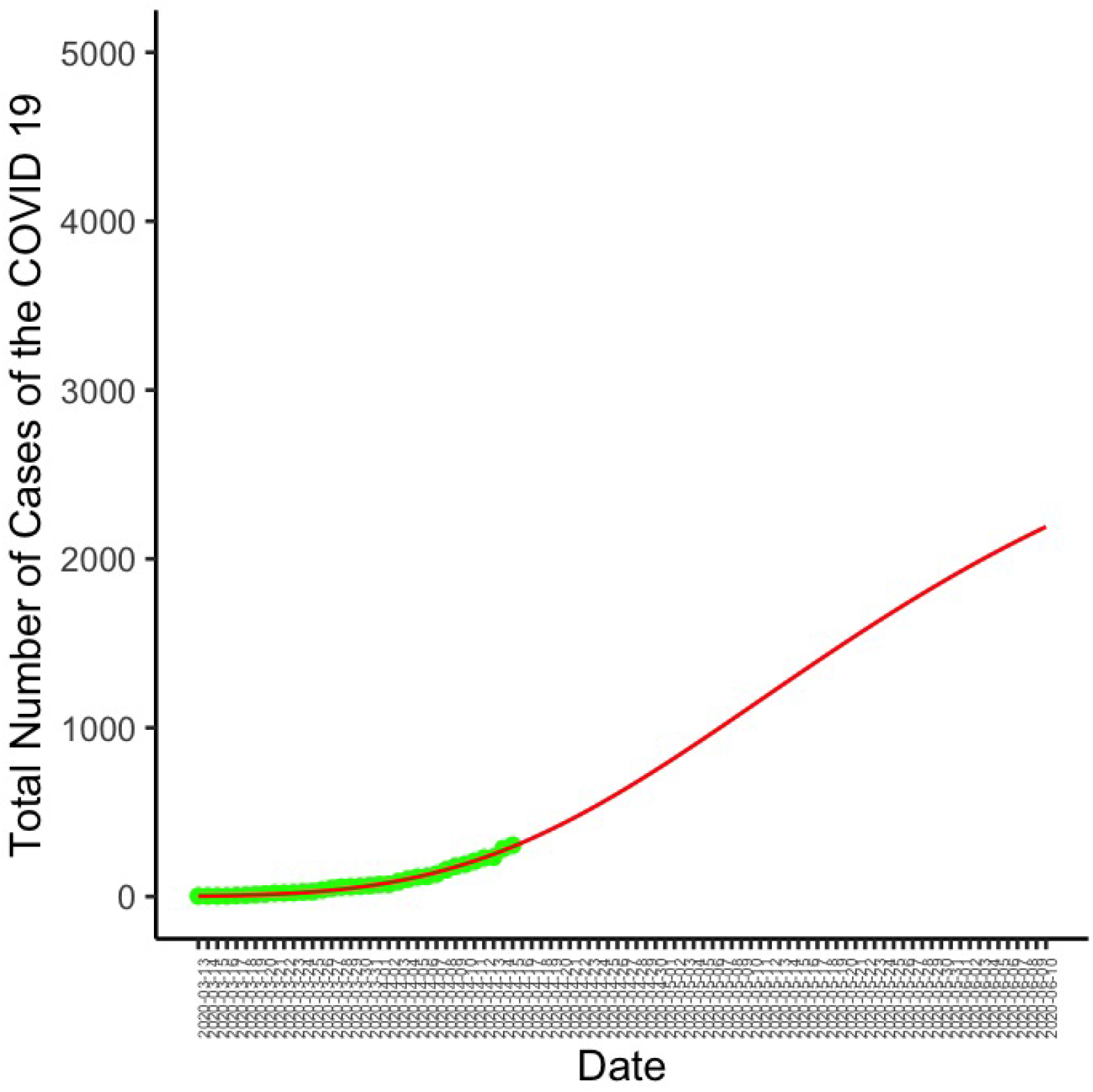
Total number of cases of COVID 19 in the State of Goias, Brazil, and fitted curve by Bayesian non-linear regression

**Figure 4:**
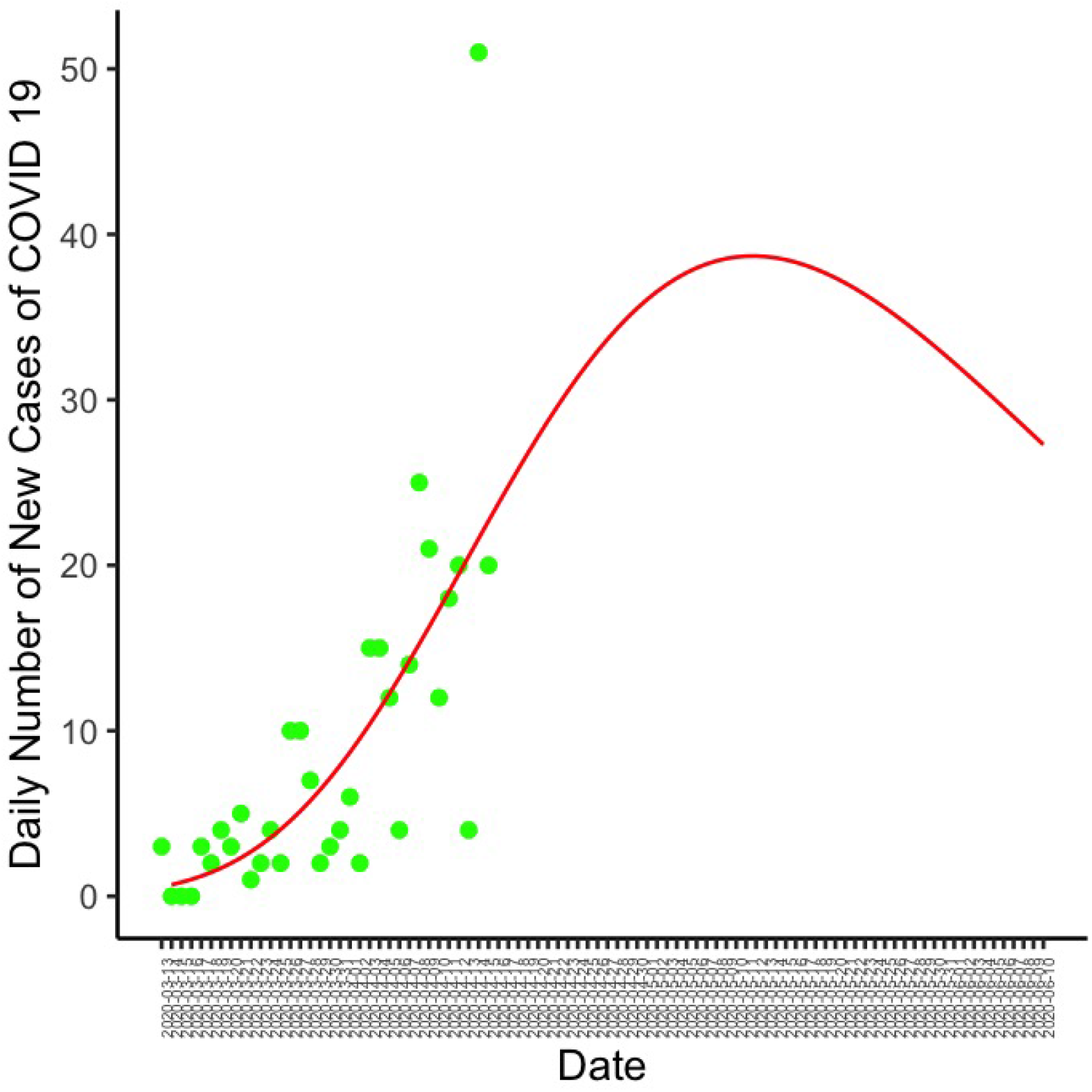
Daily number of new cases of COVID 19 in the State of Goias, Brazil, and fitted curve

## 4 Discussion

In this study, we presented a Bayesian method to predict the total number and daily new cases of the COVID 19 in the Goias state, Brazil. In this section, we are going to make comments about some technical aspects of statistical analysis and use of the results.

Firstly, why do we adopt Bayesian analysis? What are the strength and drawbacks of this sort of analysis? Researchers quite often use the compartment models to make predictions about infectious disease outbreaks ([27]; [19];[18]). This sort of model can be useful to make predictions before the outbreaks begin. However, these models do not take into account real data because they are mathematical models, frequently based on differential equations. There is no consensus in the literature about some disease information, for example, the infectious period of asymptomatic person, fatality rate. On the other hand, the frequentist statistical analysis only takes into consideration the data itself, and in our case, we have few data available for Goias state. We believe that situation leads naturally to a Bayesian statistical analysis [7]. We prefer to use information about other countries as priors, rather than only an expert opinion. Because the disease still is new, and some aspects continue no revealed.

The priors distributions based on countries’ information revealed influential. Hence, after sensibility analysis, we defined another one with higher variance, allowing that parameters also learn from the data itself.

Results from Bayesian Analysis revealed reasonable. The expected prevalence rate for Goias state is around 4.53 per 10000 inhabitants. South Korea recorded a prevalence rate equal to 2.06 until April 15, 2020. We already expect these results, considering the control measures made by both governments.

Goias is not the COVID 19 epicenter in Brazil. It is only the twelfth most populous state. For example, São Paulo and Rio de Janeiro states are more populous and have more international air traffic, which explains more cases recorded. Probably this is a reason that Goias will have a prevalence rate much less than the USA, which recorded 19.48 on April 15, 2020.

## 5 Limitations

Our model does not take into consideration changes in the disease notification politics and or quarantine or social isolation measure. So any changes made in this sense can alter the efficiency of this prediction. Also, the model ignores the sub notification problem. The target population of this study is the potential patients that can need public health facilities and resources in general. The asymptomatic and mild symptomatic are not the target. Hence, we do not recommend the use of the model to discuss the period of quarantine. However, we believe that the results of this study are enough to help the authorities in public health to make decisions.

## 6 Conclusion

We conclude that the method can be useful, and we hope this study can provide some valuable information to public health management.

## Data Availability

We used public data

## References

[1] Rodrigues A. Brazil confirms first case of coronavirus. Agencia Brasil, 26 February 2020. Available: https://agenciabrasil.ebc.com.br/en/saude/noticia/2020-02/brazil-confirms-first-case-coronavirus [Last accessed: 19 April 2020].

[2] Ali Ahmadi, Yasin Fadaei, Majid Shirani, and Fereydoon Rahmani. Modeling and forecasting trend of covid-19 epidemic in iran. medRxiv, 2020.

[3] H. Akaike. A new look at the statistical model identification. IEEE Transactions on Automatic Control, 19(6):716–723, 1974.

[4] Jessica Hwang Andrew Gelman and Aki Vehtari. Understactive information criteria for bayesian models. Statistics and Computing volume, 24:997–1016, 2014.

[5] Paul-Christian Bürkner. brms: An R package for Bayesian multilevel models using Stan. Journal of Statistical Software, 80(1):1–28, 2017.

[6] Paul-Christian Bürkner. Advanced Bayesian multilevel modeling with the R package brms. The R Journal, 10(1):395–411, 2018.

[7] Stuart G. Coles and Jonathan A. Tawn. A bayesian analysis of extreme rainfall data. Journal of the Royal Statistical Society. Series C (Applied Statistics), 45(4):463–478, 1996.

[8] Ministério da Saúde Governo do Brasil. Coronavírus brasil. https://covid.saude.gov.br, 2020 [Accessed: 2020-04-15].

[9] Edward L. Ionides Daihai He and Aaron A. King. Plug-and-play inference for disease dynamics: measles in large and small populations as a case study. Journal of The Royal Society Interface, 7, 2010.

[10] Timur V. Elzhov, Katharine M. Mullen, Andrej-Nikolai Spiess, and Benjamin M Bolker. Title R interface to the Levenberg-Marquardt nonlinear least-squares algorithm found in MINPACK, plus support for bounds, 2015.

[11] Andrew Gelman, John B. Carlin, Hal S. Stern, and Donald B. Rubin. Bayesian Data Analysis. Chapman and Hall/CRC, 2nd ed. edition, 2004.

[12] Minyoung Her. How is covid-19 affecting south korea? what is our current strategy? Disaster Medicine and Public Health Preparedness, page 1–3, 2020.

[13] Petter Holme and Naoki Masuda. The basic reproduction number as a predictor for epidemic outbreaks in temporal networks. PLOS ONE, 10(3):1–15, 03 2015.

[14] Cohen J. Scientists are racing to model the next moves of a coronavirus that’s still hard to predict. Science, 7 February 2020. Available: https://www.sciencemag.org/news/2020/02/scientists-are-racing-model-next-moves-coronavirus-thats-still-hard-predict [Last accessed: 19 April 2020].

[15] Lin Jia, Kewen Li, Yu Jiang, Xin Guo, and Ting zhao. Prediction and analysis of coronavirus disease 2019, 2020.

[16] D. Katz, S. P. Azen, and A. Schumitzky. Bayesian approach to the analysis of nonlinear models: Implementation and evaluation. Biometrics, 37(1):137–142, 1981.

[17] William Ogilvy Kermack and Anderson Gray McKendrick. A contribution to the mathematical theory of epidemics. Proceedings of the Royal Society of London A, 115:700–721, 1927.

[18] Shou-Li Li, Ottar N. Bjørnstad, Matthew J. Ferrari, Riley Mummah, Michael C. Runge, Christopher J. Fonnesbeck, Michael J. Tildesley, William J. M. Probert, and Katriona Shea. Essential information: Uncertainty and optimal control of ebola outbreaks. Proceedings of the National Academy of Sciences, 114(22):5659–5664, 2017.

[19] Benjamin F. Maier and Dirk Brockmann. Effective containment explains subexponential growth in recent confirmed covid-19 cases in china. Science, 2020.

[20] The World Health Organization. Coronavirus disease 2019 (COVID-19) Situation Report – 51. Technical report, The World Health Organization (WHO), 03 2020.

[21] R Core Team. R: A Language and Environment for Statistical Computing. R Foundation for Statistical Computing, Vienna, Austria, 2019.

[22] Tarcisio M Rocha Filho, Fabiana S. Ganem dos Santos, Victor B Gomes, Thiago A.H. Rocha, Julio H.R. Croda, Walter M Ramalho, and Wildo N Araujo. Expected impact of covid-19 outbreak in a major metropolitan area in brazil. medRxiv, 2020.

[23] Grzegorz Rzakowski, Iwona Glazewska, and Katarzyna Sawinska. The gompertz function and its applications in management. Foundations of Management, 7(1):185–190, 2015.

[24] Kathleen M. C. TjÃžrve and Even TjÃžrve. The use of gompertz models in growth analyses, and new gompertz-model approach: An addition to the unified-richards family. PLOS ONE, 12(6):1–17, 06 2017.

[25] S. Vieira and R. Hoffmann. Comparison of the logistic and the gompertz growth functions considering additive and multiplicative error terms. Journal of the Royal Statistical Society. Series C (Applied Statistics), 26(2):143–148, 1977.

[26] Worldometer. Worldometer - real time world statistics. https://www.worldometers.info/coronavirus/, 2020 [Accessed: 2020-04-15].

[27] Ya-Kui Xue Gui-Quan Sun Zhen Jin Zhi-Qiang Xia, Juan Zhang. Modeling the transmission of middle east respirator syndrome corona virus in the republic of korea. PLoS One, pages 700–721, 2015.

